# Using WhatsApp to improve self-care management among type 2 diabetes patients in Ethiopia: A qualitative study

**DOI:** 10.64898/2025.12.29.25343156

**Authors:** Helene Foss, Linn Cecilie Erlandsen, Anita Øgård-Repål, Mariann Fossum, Kalayou Berhe

**Author notes:** Corresponding author (HF).

## Abstract

**Background:** The prevalence of diabetes in low- and middle-income countries is rising, and the most important treatment is maintaining a healthy lifestyle. Good diabetes self-care management is associated with better outcomes, but barriers to adhering to its management are knowledge deficits and a lack of social support. Traditionally, diabetes self-care management education and support are conducted by health care workers (HCWs), but limited access to HCWs restricts this activity. Digital health interventions can help overcome some barriers.

**Objective:** To describe type 2 diabetic patients’ perception of using WhatsApp for diabetes education, as well as the barriers and enablers they experienced in their usage, in the Tigray region of Ethiopia.

**Method:** This study is a collaboration between researchers from Norway and a researcher from Ethiopia. A qualitative explorative and descriptive approach was adopted. The co-researcher in Ethiopia recruited the participants, and research assistants conducted 17 interviews with a semi-structured interview guide based on the technology acceptance model. The interviews were conducted in Tigrinya, transcribed and translated to English, and de-identified before analysis. The data were analysed using NVIVO 14 with reflexive thematic analysis.

**Results:** From the data, the following four themes were developed: experienced enhanced self-care, digital access to HCWs, digital support, and barriers and enablers. The participants perceived WhatsApp as highly useful. The participants said that they gained new knowledge and experienced social support and increased access to HCWs when using WhatsApp.

**Conclusion:** WhatsApp was perceived as easy to use, but some barriers were reported. This study indicates that WhatsApp may contribute to enhancing access to diabetes self-care management education and support.

## 1. Introduction

Type 2 diabetes is the predominant variant of diabetes, typically manifesting after the age of 40 and becoming increasingly prevalent with advancing age [1]. It is characterised by hyperglycaemia, insulin resistance, and relative insulin deficiency[2]. In 2024, the global prevalence of diabetes among people aged 20–79 was 11.1% (589 million), projected to rise to 13% (853 million) by 2050. In Sub-Saharan Africa, where over 90% of diabetes cases are type 2, the prevalence is expected to increase from 5% (25 million) to 5.9% (60 million) in the same period, an estimated 142% rise. In Ethiopia, the prevalence was 4.0% (2.3 million) in 2024.

One of the most important ways to treat diabetes is maintaining a healthy lifestyle with the aim to avoid or reduce acute or chronic diabetes complications [3], and patients who practice sufficient self-care management have shown better outcomes [4]. Self-care management is control and responsibility and the knowledge, beliefs, skills, and abilities to manage chronic conditions or engage in healthy behaviours [5]. Components of self-care management include a healthy diet, regular exercise, blood sugar monitoring, prescribed medication intake, and foot care [2, 6].

Self-care management has been found to be positively correlated with good glycaemic control, reduction of complications, and improvement in quality of life for diabetes patients [7]. Two systematic reviews found that more than half of diabetic patients in Ethiopia had poor diabetes self-care and that the majority of patients can significantly reduce the risk of complications by improving self-care [2, 6]. Diabetes self-care management education and support (DSMES) provide diabetic patients with the knowledge, skills, and confidence to manage their condition. DSMES is often face-to-face sessions, which can make access challenging due to remote geographical locations, transportation limitations, time constraints, and the lack of health care workers (HCWs). To overcome this and improve access to DSMES, an online delivery model may be a promising solution [7].

Digital health refers to the use of digital, mobile, and information and communication technologies in health, including both mHealth and eHealth [8]. mHealth tools have great potential to empower patients’ self-care management through information [9]. In Ethiopia, there is a shortage of educated HCWs and health facilities in providing quality diabetes care; therefore, alternative methods to educate patients must be utilised [10]. Studies in Ethiopia identified four main challenges regarding diabetes self-manangement: cultural values and beliefs, relationships and social factors, educational factors, and economic conditions [11, 12]. Furthermore, they found inadequate knowledge of self-management, such as the types of foods to eat, active lifestyles, medication adherence, and foot care [11].

In addition, they reported that social events, such as weddings and holidays, negatively influenced dietary practices and misconceptions about diabetes, and the lack of awareness made participants feel their food choices were limited. This made it difficult to follow advice on healthy diets [11]. Having social support from partners has been shown to be statistically significantly associated with good self-care practices [13]. Similarly, a study in Ethiopia revealed that patients lack social support [2]. Berhanu et al. showed that patients had severe lack of education on topics like healthy diet, blood sugar self-monitoring, physical activity, and the prevention, detection, and management of diabetes complications [14]. Other studies confirm the need for education for better understanding and compliance with self-care practices in adult diabetic patients [13, 15].

Globally, health systems are battling with exhausted HCWs and budgeting constraints [16]. In 2023, a study showed that these issues arise from community HCWs having low job satisfaction and motivation, inadequate pay, limited education and career advancement opportunities, high workloads, difficult work environments and limited supportive supervision [17]. In the Tigray region, the 2020–2022 civil war systematically destroyed much of the health care and education facilities and killed approximately 600,000 people [18], leaving the region in ruins. Currently, there is a lack of educated HCWs and facilities and a need to find alternative ways to educate patients. Digital health inventions and telemedicine can help overcome challenges like the lack of access to information and communication between patients and HCWs [19], which will, in turn, create an efficient use of HCWs’ time and reduce the impact of workforce shortages [10].

The implementation of telemedicine and mHealth depends on accessibility to mobile phones, electricity, and internet. Previous studies have shown that many Ethiopians have mobile devices and smartphones, and access to electricity and internet is good in urban areas [20]. Their willingness to use mHealth applications were associated with factors such as place of residence, age, the time it takes to reach health facilities, diabetes education level, attitude, perceived ease of use and usefulness, satisfaction with HCWs, and willingness to answer unknown calls [21, 22].

Previous studies have mainly focused on studying the quantitative outcomes of using telemedicine, with no insights into personal experience [7, 10], and Demeke [10] recommended future research exploring patients’ experiences with digital health solutions. Thus, this study aims to explore how type 2 diabetic patients perceive the use of WhatsApp-based group education for improving self-care management and the barriers they experience.

## 2. Methods

This project was conducted in a cooperation between the University of Agder, Norway, and Mekelle University, Ethiopia, and a written formal agreement was signed by both parties. The researcher with assistants from Ethiopia conducted, transcribed, and translated the interviews, and the Norwegian researchers analysed the data and wrote the manuscript. Throughout the process, the project was regularly discussed between both parties.

### 2.1 Study design

A descriptive qualitative study design was employed to explore type 2 diabetic patents’ experiences of using WhatsApp group diabetes education intervention. The study design is flexible, capable of adjusting and facilitating an in-depth and holistic understanding and, therefore, requires researchers to be involved and reflexive [23]. A descriptive qualitative design can help informants express a wide range of meanings, feelings, and behaviours [23], and the goal was to obtain an understanding of the experience of using WhatsApp.

### 2.2 Study period and research setting

The study was conducted from September 2024 to June 2025. The interviews were conducted in February 2025. The study was done in a large city in the Tigray region. In 2020, there were two referral hospitals, 14 general hospitals, and 24 primary hospitals in the Tigray region [24]. However, by the end of the two-year civil war in the region in 2022, the well-functional health care system was destroyed, and many health care students put their studies on hold. The conflict is said to be one of the deadliest conflicts in the world in recent years, which left Tigray in a humanitarian catastrophe. Infrastructure like health care and the educational system was systematically destroyed during the war [18, 25]. Overall, less than 4% of all health care facilities across Tigray were fully functional as of June 2021 [24].

The health care services provided in the Tigray hospitals for diabetic patients include follow-up visits for medical checks or clinical evaluation, counselling, medication refill, lab tests, blood pressure and weight, data records, and referral services to specialised clinics for patients with chronic complications. Individual diabetes education is provided when there is a treatment change or if the participant needs additional information on their self-care practices. In 2015, Weledegebriel [4] described the practice of providing education in a referral hospital in Tigray region as inconsistent, and it is expected that the situation post-war has gone from bad to worse.

### 2.3 Participants and recruitment

Our participants were recruited from a study conducted in winter 2024/2025 by researchers from Mekelle University, which included a group of type 2 diabetes patients. They were invited to join a WhatsApp-based group for diabetes self-management education and support (DSMES), and educational content was prepared on coping self-efficacy, attitude, and social support. Participants were added to a WhatsApp group created by the research team so they could receive the materials, which are taught in online classes and discuss self-care practices. The participants in this study formed the target group for our qualitative study, and the researchers in Mekelle recruited patients for post-intervention interviews using a purposive sampling approach [23].

The inclusion criteria were adults (>18 years) with T2DM who had a follow-up at an outpatient clinic for at least three months duration since the diagnosis of diabetes attended the WhatsApp diabetes education programme, has a smartphone, and is literate, preferably also in English. Exclusion criteria were pregnant women and critically ill or cognitively impaired patients. All the participants were selected for the interviews based on the eligibility criteria, all participation was voluntary, and no one received any incentive to be interviewed or participate in the study. Of the 20 key informants approached for interviews, 17 took part in the study.

### 2.4 Interview guide

Our research team developed a flexible semi-structured interview guide, allowing the inclusion of new topics in subsequent interviews. This interview guide was based on the technology acceptance model (TAM) to ensure that we obtain data related to our research question. The interview guide was translated from English to Tigrinya (the local language) by the local research team, and the interviews were conducted in the local language. We developed a rather strict interview guide to ensure the quality of the data. We also developed an information letter and obtained written consents to participate in the study.

### 2.5 Dataset generation

The face-to-face semi-structured interviews were conducted individually by the research assistants, and the interviews were conducted at the participants’ convenience within the hospital compound in a quiet, private, comfortable, and neutral room. The interviews were conducted between 19. February 2025 and 1. March 2025. Before the interviews, demographic data and verbal and written informed consent were obtained. Each interview was recorded, transcribed, and translated into English by the research team. All audiotaped interviews were transcribed verbatim and translated into English. The dataset was anonymised before being shared with the Norwegian team.

### 2.6 Data analysis

The interviews were transcribed verbatim in Tigrinya and translated into English by the research assistants. All transcripts were assigned a code number to guarantee confidentiality, and no personal information was shared between the two countries. Braun and Clarke’s reflexive thematic analysis was used to identify, analyse, and report themes within the local context, and analytic themes were then discussed against the TAM. We conducted a combination of inductive and deductive approaches [26].

### 2.7 Ethical declaration

The study has been conducted in accordance with the Declaration of Helsinki [27]. Ethical approval to conduct the study was obtained from the Institutional Review Board of Mekelle University, the Norwegian Agency for Shared Services in Education and Research, and the Research Ethics Committee at the University of Agder. In addition, an official letter of permission was obtained from the chief clinical director and internal medicine department at the hospital where the participants were recruited.

## 3. Results

This section is divided into four subsections, representing the four themes that present the main findings of the analysis, with associated sub-themes and illustrative quotes from the participants (Table 1). To maintain confidentiality, the term ‘IDI’ is used to refer to ‘participant’ in the order of the interviews. In our study, we had 17 participants with a mean age of 56.35 years and a mean duration of diabetes of

**Table 1.**
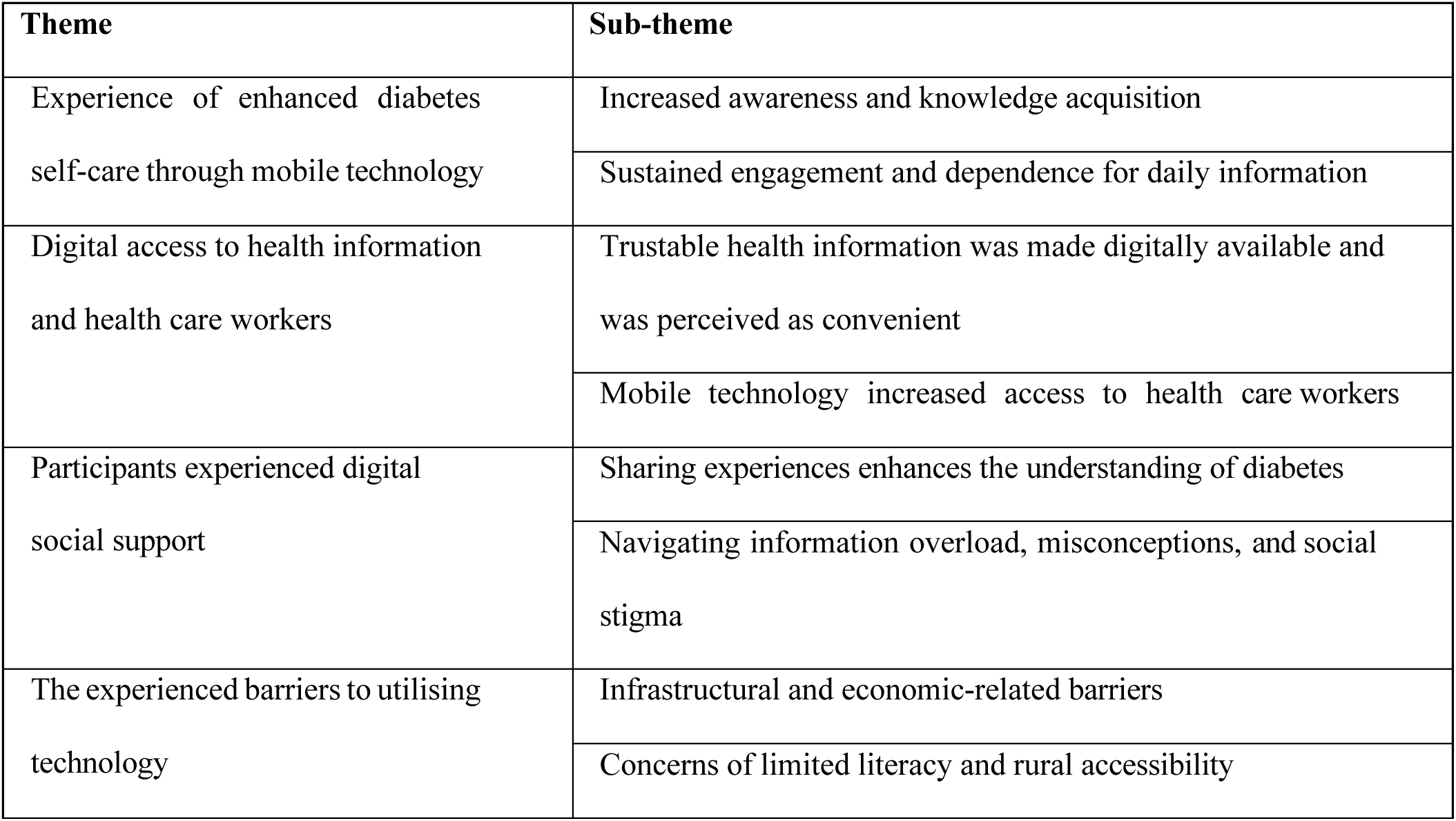
Themes and sub-themes.

| Theme | Sub-theme |
| --- | --- |
| Experience of enhanced diabetes self-care through mobile technology | Increased awareness and knowledge acquisition |
|  | Sustained engagement and dependence for daily information |
| Digital access to health information and health care workers | Trustable health information was made digitally available and was perceived as convenient |
|  | Mobile technology increased access to health care workers |
| Participants experienced digital social support | Sharing experiences enhances the understanding of diabetes |
|  | Navigating information overload, misconceptions, and social stigma |
| The experienced barriers to utilising technology | Infrastructural and economic-related barriers |
|  | Concerns of limited literacy and rural accessibility |

8.41 years. There were 12 male and five female participants. One of the participants did not have access to a smartphone, but they got hold of one through this project. Only six of the participants reported consistent access to the internet; the others visited hotels for access or used the internet at their workplaces.

### 3.1 Experience of enhanced diabetes self-care management through mobile technology

#### 3.1.1 Increased awareness and knowledge acquisition

The participants’ perception of increased knowledge was identified as a key aspect. The WhatsApp group provided them with practical insights to support improved self-care. Some had good knowledge before the intervention, while others were much more unaware of their condition and how to manage it properly. One participant said:

> *But to be frank, the information being provided after diagnosis is minimal. In my dietary management, I was careless. (IDI5)*

The new knowledge and experience the participants had gained through the WhatsApp group made them realise the important connection between diet and exercise. A few participants shared that they previously had misconceptions believing that diabetes could be cured through various homemade remedies. One participant said:

> *The influence it makes is, it changes your original thought. Hence, it changes many aspects of my behaviour. In selecting appropriate diet and doing exercises, WhatsApp-based education has huge influence. (IDI2)*

To obtain maximum benefit from the newly acquired knowledge, the next step would be to put the new knowledge into practical lifestyle changes, which some participants found challenging.

#### 3.1.2 Sustained engagement and dependence for daily information

Most of the participants were familiar with WhatsApp and had been using it for personal purposes, like keeping in touch with relatives and friends, which was seen as a benefit related to the application’s ease of use. However, the participants had not been using it for diabetes-related education prior to the intervention.

> *Since it was my first time, I was a little confused at first, but when you showed us how to use WhatsApp, I found it to be simple. We received excellent teaching. (IDI13)*

All the participants had access to the WhatsApp group, the educational content, and the group chat. Many used it regularly – both the educational content and the group chat – making calls to group members and HCWs.

> *WhatsApp-based education has significantly changed our attitude. We are, thus, introducing it to society. Therefore, we recommend that this WhatsApp-based education should be strengthened and provided continuously. (IDI2)*

The participants hoped the application would continue to be used and envisioned continuing to use the app in the future. It was suggested to increase the number of video presentations and that videos should reach a broader audience, including individuals with reading difficulties or other challenges. The participants had an overall positive attitude towards using WhatsApp in their T2DM self-care management both regarding the perceived ease of use of the technology – based on the easy access to the application and content, ease of app navigation, and an experience of simple and straightforward instructions – and the possibilities it can provide if the application is developed to a larger scale and reaches a bigger audience.

### 3.2 Digital access to health information and health care workers

#### 3.2.1 Trustable health information was made digitally available and was perceived as convenient

The application was perceived as user-friendly, with content readily available always. All participants perceived the health information provided by professionals through WhatsApp as credible and easy to understand. A key factor contributing to this was the use of the participants’ native languages. Although some experienced challenges with technical and professional terms that lack direct translations in Tigrinya, support from group members and health care professionals helped mitigate these challenges, with prompt explanations and clarifications. Some participants attributed their confidence in the information to the presence of scientific explanations and practical solutions accompanying the guidance provided.

> *WhatsApp is with you. The brochures and the manual are there, and whether there is internet or not, you just open your WhatsApp and read at any time you need. (IDI5)*

The availability of multiple content formats was perceived as beneficial, as individuals have different learning preferences. Several participants emphasised the value of video materials as a more effective learning experience compared to text-based content. Some participants noted that listening to speech and watching videos was preferable to reading, particularly for individuals with reduced vision or limited literacy, enabling them to communicate effectively. WhatsApp was perceived as an appropriate tool for diabetes self-care management, and a significant advantage of the application is its ability to provide continuous access to health information without requiring physical attendance at meetings. WhatsApp reduced the need for travel to clinics, thereby decreasing the cost of time, money, and other related expenses. Some participants also saw it as an advantage for HCWs, who can serve more people at once. Participants concurred that WhatsApp was highly convenient, being accessible everywhere as long as there is internet access or the content has been downloaded already. It was timesaving, allowing them to obtain information whenever needed and receive prompt responses without the necessity of waiting for and traveling to hospital appointments.

#### 3.2.2 Mobile technology increased access to health care workers

Having access to HCWs was perceived as a significant benefit of the WhatsApp group education programme. One participant noted that the WhatsApp programme’s value lies in the active interaction between HCWs and group members. This access enabled participants to ask questions and receive prompt answers even from home without requiring hospital visits.

> *You get necessary information daily there, and you can’t go to health institutions daily; you can ask whatever you from where you are. (IDI1)*

Members of the WhatsApp group expressed gratitude for having access to doctors to ask questions at their convenience. Although the exact frequency of these interactions is unknown, responses indicated that members frequently reached out to HCWs in the group, with some doing so daily. Questions were posed both in group chats and as direct messages to HCWs.

### 3.3 Participants experienced digital social support

#### 3.3.1 Sharing experiences enhances the understanding of diabetes

The participants strongly appreciated being included in the WhatsApp group and the experience of digital social support, which some had been missing before the intervention. Participants described how they have come to know some of the other members, fostering an underlying sense of being part of a community. Sharing experiences was highly valued and contributed to a deeper understanding of the illness and to promoting physical exercise and dietary management.

> *Since diabetes affects everyone differently, it was beneficial for us to share our experiences in the group. Thus, it was really beneficial to engage with other diabetic patients and group members. (IDI17)*

The members of the group shared information and knowledge among themselves as well as with family, friends, and neighbours. This impacts how their families support them by making better food-related choices. Participants described how they actively promoted the WhatsApp group to others in the community, spreading knowledge outside the group. Some participants mentioned misconceptions commonly spread on other social media platforms and in the community.

> *In general, starting from our family members, we are introducing diabetic information and this method of education among the community. (IDI2)*

#### 3.3.2 Navigating information overload and social stigma

Some participants felt overwhelmed by some extreme and repeated information. For instance, repeated discussions about complications, such as amputations, induced fear among some members. This fear was compounded by the fact that many participants had previously obtained their information about diabetes from YouTube or Facebook, leading to scepticism about the shared information.

> *It is good to know that the disease has complications, but when it becomes extreme and repeated too much, I sometimes say that this should be stopped. (IDI9)*

Additionally, participants discussed the social stigma associated with diabetes. Initially, some hesitated disclosing their condition but became more open after joining this group. The sense of community and support within the group helped alleviate some of the stigma, encouraged more open discussions about their condition, and helped them reconcile with it.

> *I was ashamed of it (diabetes). But now I have joined WhatsApp, and there is nothing worrying about it. (IDI5)*

### 3.4 Experienced barriers to utilising technology

#### 3.4.1 Infrastructural and economic-related barriers

Network availability and power outages were challenges for many of the participants. While some of them reported having internet access at home, the majority did not, requiring them to visit hotels, cafes, or workplaces to get and maintain access. Some also highlighted the difficulties related to battery capacity and charging their devices.

> *There is no consistent supply. There is an interruption of electric power (…), there is also an interruption of internet service. (IDI4)*

Economic limitations were frequently mentioned, as many participants could not afford mobile data. One participant stated:

> *The challenge I faced is limitation of internet use due to economic problem. I tried to deal with it by going to Wi-Fi centres and using mobile data when I got money. (IDI5)*

#### 3.4.2 Concerns of limited literacy and rural accessibility

Some participants emphasised that technology was not available to everyone, particularly individuals in rural areas and those with limited literacy. The most prominent suggestion to addressing this challenge was the need to extend the reach of the WhatsApp intervention to more remote and rural areas in Ethiopia.

> *Its coverage and accessibility should be considered because there are many individuals at urban and rural settings who are illiterate and unable to operate mobile phones. (IDI 9)*

## 4. Discussion

This study aimed to describe the experiences of how type 2 diabetes patients perceived using WhatsApp for diabetes education and the barriers and enablers they experienced in their usage. The presentation of our results is based on the TAM [28], which focuses on perceived ease of use and perceived usefulness, and we added novel factors such as social factors, access to the internet and power, support and access to HCWs, and barriers and enablers to utilising this intervention. Therefore, we added the Individual and Family Self-Management Theory IFSM [5] to capture these factors.

The main findings of this study are discussed in relation to theory and previous studies. The main themes are experiences of enhanced diabetes self-care management through mobile technology, digital access to health information and HCWs, participants’ experiences of digital support, and the barriers and enablers to utilising technology, which is consistent with themes in Firdisa et.al [22] study. All participants in our study stated that they had increased their knowledge and discovered new and valuable information through the WhatsApp group education programme. Moreover, many participants in our study said that they had little information on self-care management and had poor self-care management before joining the WhatsApp group. Ketema et al. [22] also concluded that more than half of diabetic patients in Ethiopia had poor dietary practice, and recommends intervention programs should focus on improving the knowledge level to improve the self-care practice of diabetic patients. This is consistent with research by Firdisa et al. [22] and Ketema et al. [6]. Moreover, a study by Weledegebriel [4] stated that education given in the outpatient clinic is inconsistent, and barriers to self-care management in the Tigray region in Ethiopia were a lack of education and insufficient knowledge [2].

In this study, the participants reported gaining new and valuable knowledge on self-care management through the WhatsApp training. Additionally, the lack of HCWs restricts the availability of the traditional education settings, but from our study, using WhatsApp for this purpose is beneficial. A study in Ethiopia by Berhanu et al. showed that there was a severe lack of competence among HCWs in providing health education and a lack of regular, structured diabetes education. However, after the study’s intervention, there was a remarkable change related to the availability of educational materials and the involvement of HCWs, which also led to increased knowledge among the patients [14].

On the other hand, to be able to utilise the technology, users must be willing to use it. Previous studies from Ethiopia reveal a high rate of willingness, and this is related to, among others, perceived ease of use and usefulness. Patients who perceived the technology as a tool to improve self-management also increased their intention to use it [21]. At the same time, if a system is perceived to be difficult or time-consuming to use, the application’s usage will be weighed by the effort required to use it [29]. This is also supported by Greve et al. [30], who highlighted the importance of the application having a clear purpose, feeling like a contribution, and providing an advantage for the user [30].

Our study shows that the participants acknowledged self-care management components and found them all important for managing their diabetes after the education they received on WhatsApp. This supports the study of Aminuddin et al. [31], which indicated that participants who received and participated in such interventions had better self-efficacy, self-care activities, and health-relevant outcomes, and thus, the intervention was considered beneficial [31].

As described earlier, and supported by Ryan and Sawin [5], self-management refers to behaviours to manage chronic conditions and enhance health behaviours and includes the individual’s abilities and self-regulation skills, knowledge and beliefs, and the social facilitation and enhancement of these processes. The goal is to manage healthy routines that aim to prevent or lessen illness [5]. Among the barriers described by Berhe [2] were challenges related to adapting to a new lifestyle and changing old habits in line with new knowledge. In this study, some participants noted that they experienced increased knowledge after the intervention but had challenges adopting the knowledge into practical use and lifestyle changes. Similarly, the IFSM theory describes this challenge in the process dimension – the ability to change health behaviours is dependent on the enhancement of knowledge and beliefs, engagement in changing health behaviours, expectancy of outcomes, and socioeconomic status. Moreover, a study by Ryan et al. revealed that patients are largely dependent on the social support, influence, and collaboration between the participants, their families, and the HCWs [5].

Most of the participants in our study were familiar with WhatsApp and had been using it before the intervention, mostly to keep in contact with family and friends. According to Walle et al. [21], the user’s willingness to use mHealth solutions is crucial to adopting mHealth interventions. We found that most of the participants had a positive attitude and willingness related to the use of the application, which they found easy to use, available, and accessible. This is consistent with a study by Marikyan et al. [32], which mentioned that users’ attitudes towards using a system is strongly connected to the users’ perceived usefulness and ease of use of the system, as described earlier [32]. Moreover, in Walle et al.’s study [21], most of the participants were willing to use mHealth applications and found them useful, which is also confirmed by Firdisa et al. [22]. They found that factors such as place of residence, age, mobile device ownership, attitude, perceived ease of use, and usefulness, among others, affected participants’ willingness to use the applications. Similarly, a study by Walle et al. [21] showed that participants under the age of 30 were more willing to use mHealth applications than older participants. Nevertheless, the participants in our study ranged from 39 to 75 years old, and most of them were willing and positive towards using WhatsApp and intended to continue using it for this purpose. Moreover, in Walle et al.’s study [21], the willingness and the intention to use the applications were strongly connected to the participants’ attitude towards it; if the participants find the application useful and believe it will benefit them, they are more likely to use it [21].

All participants perceived health information from HCWs as credible. Many had previously relied on platforms like YouTube and Facebook, where information, especially from non-professionals, was often seen as unreliable. In contrast, participants expressed greater trust when HCWs provided scientific explanations. Consistent with this, Amanu et al. [33] found that HCWs and television broadcasts were widely used sources of health information among their participants. Moreover, a study by Mengiste et al. found that the most common source of diabetes-related information was HCWs, followed by mass media, the internet, family, friends, magazines, and newspapers [34]. We acknowledge that participants seek information online and on social media, but there is limited or no control over the reliability of the content. In this intervention, the reliability of the content is ensured by HCWs and their teams.

Participants highlighted the value of being able to revisit educational content at their convenience, even offline after downloading. Videos were perceived as particularly effective, and key messages delivered through voice recordings enhanced accessibility. This underscores the importance of providing available, understandable, and trustworthy health information. Similarly, the Be Healthy Be Mobile initiative emphasised that it is important to consider language, tone, clarity, health literacy, and technological literacy when writing messages [8]. Additionally, a study by Firdisa et al. stated that mHealth interventions can save time, money, and resources by reducing the need for travelling to clinics [22]. The participants receive information, access to educational content, access to group chats with the other participants, and direct access to HCWs through the WhatsApp-based education programme. Additionally, they stated that the WhatsApp-based education programme could reach individuals in more rural areas. In line with this, a study by Correia et al. showed that living in remote areas, lack of transportation, and time constraints can restrict people from accessing health resources, and telemedicine can help overcome these challenges [19]. Several participants reported receiving limited information during brief check-up visits, often spaced months apart. They noted a lack of access to HCWs between appointments, which contributed to feelings of insufficient support and information. The SOWN 2020 report estimated a current nursing shortfall of 5.3 million in low- and middle-income countries, and the GBD analysis from 2019 showed that Sub-Saharan and North Africa had the lowest availability of human resources for health [16]. In Ethiopia, the ratio of doctors, nurses, and midwives’ to the population is 0.8:1000, which is less than the World Health Organization’s minimum recommendation level of 4.45:1000 [10]. Participants felt that WhatsApp-based group education improved access to HCWs. Many frequently contacted HCWs via direct messages, texts, or voice notes, especially when challenges arose. According to Correia et al. [19], digital health inventions can improve communication between HCWs and patients [19].

One key benefit of the WhatsApp group was social support. Participants encouraged each other in physical activity and diet and described a sense of connection that extended beyond the digital space. This is supported by Brew-Sam [35], who identified the patients’ social networks as an important factor for empowerment and self-management supported by technology [35].

In addition to the in-group support, the information was shared outside the group with family, friends, and neighbours. This way, the intervention not only impacts the participants directly but also increases the knowledge in the community, thereby also reducing the negative impacts from others. In light of the IFSM theory, the experience of unity and social support in the WhatsApp group can give HCWs a more comprehensive perspective of the focus on each patient, taking into account the importance of social and community networks and the inclusion of family and friends [5].

Several participants had previously used Facebook and YouTube to obtain information related to diabetes self-care management, and some misconceptions were discussed and clarified in the group, such as whether diabetes is curable in other parts of the world and whether honey or alcohol is beneficial for diabetes patients. In relation to this, Desse et al.’s study stated that the sociocultural context influenced beliefs and interpretations, and in his study, they identified a belief that medications for diabetes are harmful, with advice to use herbs, rituals, and holy water in self-management [11]. Some participants previously hesitated disclosing their condition due to stigma. Through group support and increased knowledge, they gained confidence, challenged misconceptions, such as diabetes being a result of sin, and became more open, helping to spread awareness. This can also be seen in light of Desse et al.’s study [11], where cultural beliefs and values and social factors are challenges to the self-management of diabetes.

Network availability and power disruptions were identified as challenges for many of the participants. This is consistent with Firdisa et al.’s study [22], which identified infrastructure-related barriers like the ones presented as major barriers to utilising technology in Ethiopia. Most of the participants in our study did not have internet access at home, requiring them to use the internet at their workplaces or visit hotels or cafes. Similarly, in 2024, Biga et al. identified unequal access to broadband as a barrier to accessing web-based patient portals, and patients with restricted ability to access broadband developed fewer skills in utilising technology [36]. In our study, 16 of the 17 participants had a smartphone. This finding is in line with Firdisa et al. [22], Jemere et al. [37], and Walle et al. [21], who found that many individuals in Ethiopia have access to a mobile phone or smartphone, and an accessible application like WhatsApp seems to be a suitable solution to save money and time for the participants, HCWs, and health care services [38]. Moreover, in Nigeria, a high mobile phone usage was found to be a key driver of telemedicine [39], and having access to a smartphone is a prerequisite for utilising this technology [21]. Some participants faced economic constraints and family responsibilities, making it difficult to prioritise health education. Limited access to mobile data was a common barrier, which some overcame by using Wi-Fi in public places like cafes and hotels. This is supported by the findings of Shiferaw et al. [40] and Firdisa et al. [22], demonstrating that economic status and monthly salary were a major barrier to utilising technology, as well as not having a mobile phone. Another identified barrier with technology was power outages, in this case, related to charging of devices and battery capacity on mobile phones. In 2016, numbers showed that 93% of urban households had access to electricity, compared to only 8% of rural households; however, interruptions in electricity supply were a challenge and could last from hours to days [20]. In our study, the participants mentioned that they could charge their devices at their workplaces or other suitable locations. If patients have no access to power to charge batteries, they cannot use their phones or the application, which is a major infrastructural barrier to utilising the technology.

Another concern that was raised by most participants in this study was accessibility for uneducated and less digitally literate individuals in society, as well as for elderly people and those living in rural areas. According to Shiferaw et al. [40], the eHealth literacy skills for obtaining health-related information among patients in a certain hospital in Northwest Ethiopia were relatively low, but most of the patients used the internet daily or several days a week. In comparison to the study by Walle et al. [21], younger patients seemed to have higher eHealth literacy skills, and factors like educational status, residence, health status, salary, frequency of internet use, and computer literacy level, among others, were related to the patients’ level of eHealth literacy skills.

### Methodological considerations

We regularly considered and discussed the reliability of our study, the accuracy of the data collection process, and the data analysis and use. We are also aware that the reliability and validity of an interview process can be affected by the connection between the participant and the researcher. We chose to use the TAM as a framework and developed the interview guide based on it to capture the participants’ experiences related to the theoretical variables in the TAM. To prevent bias, such as participants feeling obliged to be positive about the intervention, we involved research assistants with previous experience in conducting interviews. The research assistants spoke the same language as the participants to avoid errors in interpretation and understand the participants’ communication styles, which enhanced the accuracy of the findings; the research assistants were familiar with the study’s setting, norms, and values. In addition, the study’s purpose, process, and assurance of information confidentiality were explained to the participants. To gain a deeper understanding, the participants’ information was probed further for clarity. At all times, the interviewers remained ‘experts’ in the subject and maintained a non-judgmental attitude to prevent possible bias and assumptions, thus enhancing the confirmability or neutrality of emerging issues. This was to ensure that the cultural context was well considered and to avoid misunderstandings.

### 4.1 Limitations of the study

The findings in this study are based on a limited group of participants and in a low-resource setting in an urban part of Ethiopia. This situation might have restricted transferability of the findings to other communities. Moreover, some researcher subjectivity might arise during data interpretation, related to little research experience and unfamiliar cultural context, with the risk of losing or misunderstanding information in the translation process.

## 5. Conclusion

This study revealed that type 2 diabetes patients in an urban setting in the Tigray region of Ethiopia perceived the WhatsApp-based group education programme as highly useful, as all participants gained new knowledge and experienced social support and increased access to HCWs after joining the group. Based on these factors, participants experienced the benefits of enhanced diabetes self-care management through using the WhatsApp group. In addition, the application was perceived as easy to use in terms of navigation and understanding the content. The participants were all positive about using the application for this purpose and were all active in different ways of using it. The participants addressed infrastructural and socioeconomic factors and the concerns for those with limited literacy and accessibility in rural communities as barriers and enablers to using WhatsApp and utilising technology in general to improve their T2DM self-care management. They also suggested extending the WhatsApp group to more rural communities.

## 6. Implications for further practice

We emphasise the importance of considering the cultural context. Special attention should be paid to vulnerable groups, including women and girls, older adults, people with disabilities, internally displaced persons, and those with limited literacy and digital literacy, to ensure that interventions are accessible and responsive to diverse needs. These groups often face significant barriers to accessing digital health services due to infrastructural limitations, affordability issues, and sociocultural factors.

### 6.1 Implications for further research

As many of our participants pointed out, extending the intervention to reach people in more rural areas of Ethiopia is a suggested next step in the extension of this study. Further research should examine the impact on health literacy and user experiences of other technical health-related applications, particularly given the lack of studies assessing useŕ experiences and outcomes in many implemented solutions. It would be valuable to investigate the impact of health literacy and user experiences with other health-related technological applications, particularly given the lack of studies measuring user experiences and outcomes in the majority of implementations. This could be conducted as a usability study or a quantitative study of actual use.

## Data Availability

The data underlying this study are contained in an unpublished master's thesis and are not stored in a public repository. The thesis and associated data are available upon reasonable request from the corresponding author.

